# Synovial Transcriptome Profiling for Predicting Biological Treatment Response in Rheumatoid Arthritis: A Feasibility study

**DOI:** 10.1101/2024.08.28.24312608

**Authors:** P.N. d’Ailly, O.J.M. Schäffers, C. Deugd, M.A. Versnel, H.J.G. van de Werken, E.M.J. Bindels, S.W. Tas, J. Gribnau, N.W.L. Schep, R.J. Bisoendial

**Affiliations:** Department of Surgery, Maasstad hospital, Rotterdam, the Netherlands; Department of Obstetrics and Fetal Medicine, Erasmus MC, Rotterdam, The Netherlands; Department of Developmental biology, Erasmus Medical Center Rotterdam, The Netherlands; Department of Rheumatology and Clinical immunology, Maasstad hospital Rotterdam, the Netherlands; Department of Immunology, Erasmus Medical Center Rotterdam, The Netherlands; Department of Hematology, Erasmus Medical Center Rotterdam, The Netherlands; Department of Rheumatology and Clinical immunology, Amsterdam University Medical Center location AMC, Amsterdam the Netherlands

## Abstract

**Introduction:** Disease-Modifying Anti-Rheumatic Drug (DMARD) treatment fails to achieve clinical remission in a substantial proportion of patients with rheumatoid arthritis (RA). Patient-derived synovial tissue (ST)-signatures, thought to determine this heterogeneity of treatment responses, can be studied by single-cell RNA sequencing (scRNA-seq).

**Study aims:** The first aim was to obtain viable ST from RA patients using wrist arthroscopy. The second aim was to identify patient-specific transcriptome signatures from the ST omics data that relate to clinical course and treatment responses in RA.

**Methods:** Radiocarpal and midcarpal synovectomy was performed using a standard set-up wrist arthroscopy. Single-cell suspensions of ST from affected wrists of two RA patients and a control subject were processed on the 10X Genomics Chromium Platform. Seurat was used for downstream analysis.

**Results:** In two RA patients and one non-inflammatory control, ST was successfully removed during wrist arthroscopy. No surgical complications occurred. For the RA patients and control, 17,176 and 7,884 high-quality cells were analyzed, respectively. Apart from enrichment of cell compartments in RA, including those of B- and plasma cells, T cell populations, NK cells, and macrophages, we observed interpatient variability that may influence the relationship between RA synovial signature and clinical phenotype, potentially also affecting treatment response and outcome. In-depth analysis of the prevailing cell-type abundance phenotype (CTAP) in the RA patients, as described previously, provided insights into the extent to which these CTAPs may be used to predict treatment responses.

**Conclusion:** In this feasibility study, we demonstrated that wrist arthroscopy successfully retrieves ST with good tissue viability, which may provide informative and high-quality transcriptomic data for predicting therapy response at an individual level.

## INTRODUCTION

Rheumatoid arthritis (RA) represents a common autoimmune-mediated inflammatory joint disease that frequently involves the wrist and small hand joints, and may result in progressive joint destruction and loss of function[1, 2]. First-line treatment with conventional synthetic Disease-Modifying Anti-Rheumatic Drugs (csDMARDs) fails to achieve clinical remission in a substantial proportion of RA patients, as stressed by a recent systematic review reporting on the American College of Rheumatology (ACR) response criteria ACR 20, ACR 50 and ACR 70 of only 73%, 51% and 32% respectively in DMARD naïve patients [3, 4].

Disease duration and activity, tightly bound by heterogeneity in cellular and molecular signatures prior to initiation of standard treatment modalities are thought to contribute to the variable therapy reponses and clinical evolution of RA [5, 6]. Failure to control disease activity or unintentional adverse effects demands a switch to biological (b) or targeted synthetic (ts)DMARDs, which are costly pharmaceuticals that specifically target disease-associated cytokines and inflammatory pathways. In the absence of clinical instruments that predict the individual therapy response, patients are nowadays treated empirically and sequentially with different b- or tsDMARDs according to international guidelines until an effective treatment is found. In addition to prolonged disease activity, off-target effects, long-term work absence, and higher socioeconomic costs, patients are at risk of developing a ‘refractory state’ of the inflamed joint, which in RA corresponds to a failure to respond to two or more bDMARDs[7]. The prevalence of the ‘inflammatory’ refractory subtype with objectifiable signs of arthritis, lies between 6 to 17% [7, 8]. Amongst known determinants associated with poorer disease control are delayed initiation of therapy, disease duration, and number of prior DMARDs.

Integrative omics approaches, including single cell RNA sequencing (scRNA-seq) and spatial transcriptomics, have emerged as promising instruments to interrogate the communication networks between resident and immigrant cell types within synovial tissue (ST) and synovial fluid cells. These instruments help to determine the underlying immune aberrations that drives the patient’s disease state[9–12].

The aims of this feasibility study were to obtain ST samples with excellent tissue viability from RA patients using wrist arthroscopy, and to identify patient-specific transcriptome signatures in arthroscopically derived ST that may guide the stratification of bDMARD-naïve RA patients to long-term treatment with b- or tsDMARDs.

## METHODS

### Study design

For this feasibility study, samples derive from patients who participate in the ARCTIC trial, a randomized control trial (clinicaltrials.gov NCT04755127) of which the main objective is to compare short- and long-term effects of therapeutic synovectomy versus intra-articular corticosteroid injections in terms of wrist function, pain, progression of arthritis, return to work, and overall cost-effectiveness[13]. The in- and exclusion criteria for the ARCTIC trial and the current feasibility study can be found in Table 1 and 2. Patients, who are randomized for the arthroscopy arm of the study, undergo synovectomy of the radiocarpal and midcarpal compartments of the wrist under regional anesthesia. Control samples are collected from patients with non-inflammatory conditions including ganglion cyst, post-trauma injury or osteoarthritis, who at an oppertune moment undergo joint surgery, hindering exact matching for age and gender (embedded in the LYmphatics in Psoriatic Arthritis (PsA-LY) study; NL55419.101.15).

**Table 1.** Eligibility criteria for the patient group of the ARCTIC trial.

| **Table 1.** Eligibility criteria for the patient group of the ARCTIC trial |  |
| --- | --- |
| <u>Inclusion criteria</u><br>• Age $\geq 18$ years<br>• Diagnosis of RA according to the revised 2010 ACR/EULAR Rheumatoid Arthritis Classification Criteria <u>OR</u> PsA according to the CASPAR criteria<br>• Currently experiencing an exacerbation defined as an increase in DAS28 $> 1.2$ or $> 0.6$ if DAS28 $\geq 3.2$ compared to last DAS28 measurement (maximum six months before), or an exacerbation clinically diagnosed by a rheumatologist.<br>• The exacerbation is refractory to systemic cDMARDs for at least three months, defined as 'no response' according to the EULAR response criteria (ref)<br>• Wrist arthritis, that is diagnosed clinically, is the predominant symptom | <u>Exclusion criteria</u><br>• Previous or current treatment with biological (b) DMARDs<br>• Current inflammatory joint disease other than RA or PsA (e.g. gout, reactive arthritis, Lyme disease)<br>• Subjects who are pregnant or intend to become pregnant during the study<br>• Intra-articular corticosteroids injection in the wrist in the last 3 months<br>• Previous wrist surgery<br>• Severe osteoarthritis with malformations of the wrist<br>• Congenital abnormalities of wrist function or motion |

**Table 2.** Eligibility criteria for the control group.

| **Table 2.** Eligibility criteria for the control group |  |
| --- | --- |
| <u>Inclusion criteria</u><br>• Age $\geq 18$ years<br>• Patients are undergoing arthroscopic synovectomy of the wrist for therapeutic purposes (e.g. post-traumatic wrist symptoms)<br>• Removed synovial tissue is otherwise discarded (not needed for pathologic examination or culturing) | <u>Exclusion criteria</u><br>• Patient with inflammatory joint disease, septic arthritis, a ganglion cyst of the wrist or wrist trauma less than four weeks before arthroscopy.<br>• Previous or current treatment with biological (b) DMARDs<br>• Intra-articular corticosteroids injection in the wrist in the last 3 months |

**Table 3.**
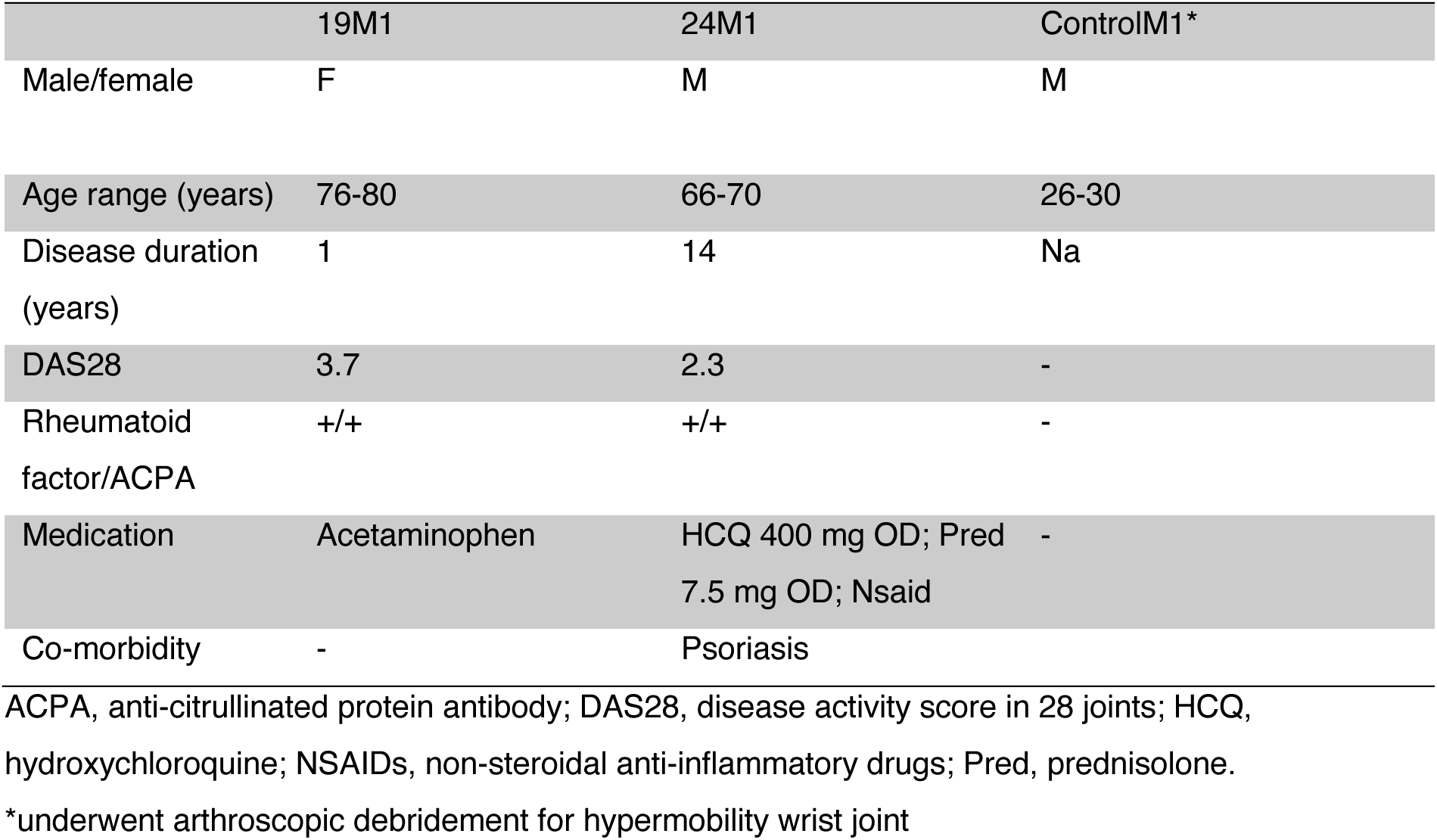
Demographic and clinical parameters subjects.

|  | 19M1 | 24M1 | ControlM1* |
| --- | --- | --- | --- |
| Male/female | F | M | M |
| Age range (years) | 76-80 | 66-70 | 26-30 |
| Disease duration (years) | 1 | 14 | Na |
| DAS28 | 3.7 | 2.3 | - |
| Rheumatoid factor/ACPA | +/+ | +/+ | - |
| Medication | Acetaminophen | HCQ 400 mg OD; Pred 7.5 mg OD; Nsaid | - |
| Co-morbidity | - | Psoriasis |  |

ACPA, anti-citrullinated protein antibody; DAS28, disease activity score in 28 joints; HCQ, hydroxychloroquine; NSAIDs, non-steroidal anti-inflammatory drugs; Pred, prednisolone.
\*underwent arthroscopic debridement for hypermobility wrist joint

### Study procedures

In the ARCTIC study, demographic and clinical parameters are prospectively obtained such as age, gender, disease duration, anti–citrullinated protein antibody (ACPA) and rheumatoid factor (RF) status, erythrocyte sedimentation rate (ESR), C-reactive protein (CRP) level, swollen and tender joint counts, global health score reflected by a visual analogue scale, use of cDMARD, and concomitant medications. All cDMARDs are paused two weeks before arthroscopy to ensure sufficient wash-out. Patients are allowed to use stable dose of systemic corticosteroids and analgesics as normal. The arthroscopic procedure is performed on outpatient basis according to international standards, by a highly dedicated hand surgeon using a standard arthroscopy setup. No postoperative restrictions are imposed. A mechanized shaver is used to remove the affected ST. Immediately after synovectomy, ST biopsies are recovered, pooled, and stored in cryovials (CryoStor CS10, BioLife solutions) in liquid nitrogen.

### Histology

ST were formaline-fixed and embedded in paraffin. Fixed tissue sections (3-5 μm) were stained with hematoxylin/eosin. For assessment of the degree of synovitis and immunophenotype, immunohistochemical staining was performed using antibodies against CD3 (T cells); CD4 (T helper cells); CD8 (cytotoxic T cells); CD68 (pan-macrophage marker); CD20 (B cells), and CD138 (plasma cells) and digitally analyzed using QuPath[14].

### Sample preparation for scRNA-seq

ST was processed as described earlier[15]. In brief, ST samples were quickly thawed, and enzymatically digested into single-cell suspensions by incubation with a mixture of DNAse I (final concentration 100mg/mL; Roche) and Liberase TL (final concentration 100µg/mL; Roche) for 30 minutes at 37°C [15]. After dead cell exclusion on a FACS ARIA (BD Biosciences), viable cells were resuspended to 700-1000 cells/µL in PBS with 0.2% BSA.

### Library preparation and sequencing

Single cell suspensions were processed on the 10X Genomic Chromium Platform using the Chromium Next GEM Single Cell 3′ Reagent kit v3.1 with Dual Index Kit (10X Genomics) according to the manufacturer’s instructions. In summary, 15,000 cells were loaded into each channel of a chip to undergo partitioning into gel beads in emulsion (GEMs). Within the GEMs, cells were lysed and subjected to barcoded reverse transcription of RNA. Subsequent steps involved breaking the GEMs, amplification, fragmentation and addition of adapter and sample index. Libraries were pooled and sequenced on an Illumina NovaSeq 6000 instrument (minimum coverage of 25,000 raw reads per cell).

### Data analysis

Raw FASTQ files were processed using the Cell Ranger software pipeline (version 7.0.0, 10X Genomics). For quality control, cells with fewer than 500 counts, 250 detected genes and low complexity (log10 value of number of genes detected per count < 0.8) were removed. Genes with zero expression in all cells and that are expressed in less than 10 cells were removed. Downstream data analyses were performed using the R package Seurat (version 5.0.1, Satija lab). Datasets were normalized by the ‘SCTransform’ function. Cell cycle heterogeneity was scored and differences between S and G2M cell cycle phases were regressed out using the functions ‘CellCycleScoring’ and ‘ScaleData’. Integration of the datasets was performed according to the Seurat alignment workflow using the Leiden algorithm[16]. UMAP analysis was performed using the ‘RunUMAP’ function including 20 principal components. Clusters were identified using the ‘FindClusters’ function with a resolution of 0.8. Cluster marker genes were identified by differential expression analysis using the ‘FindAllMarkers’ function with a minimum log fold change value > 0.25 and genes detected in a minimum of 25% cells in each cluster. Clusters were annotated using canonical markers. Differentially expressed genes (DEGs) were plotted using the EnhancedVolcano package[11]. Gene signatures were plotted using the ‘AddModuleScore’ function. Permutation tests to calculate relative proportional cluster differences were performed with the scProportionTest package (https://gitbhub.com/rpolicastro/scProportionTest). Cell communication analysis was performed with Cellchat[17], a database-driven application that in context of ligand-receptor pair interactions together with cofactors predicts cellular responses (≈defined by input (receiver) and output (sender) signals) through network analysis and patteren recognition

## RESULTS

### Demographic data and wrist arthroscopy

For this feasibility study, we selected two patients that were seropositive for rheumafactor and anti-citrullinated protein antibody (ACPA) and presented with a monoartritis of the wrist, as well as one non-inflamed control. In all three cases, the affected ST was successfully identified and removed during wrist arthroscopy. No surgical complications occurred.

### Unsupervised clustering of scRNA-seq data

Synovial samples were collected and processed to prepare scRNAseq datasets (Figure 1A). After quality control, we obtained 25,060 cells from the three samples combined. To dissect the relative composition of the ST, we integrated all datasets, and performed graph-based clustering. Using canonical markers [10, 15, 18], we could identify 25 cell clusters, including 5 different subsets of fibroblasts, 5 types of endothelial cells, a population of cycling cells, myeloid populations comprised of 4 types of macrophages and dendritic cells, a stromal/mural cell compartment, and effector populations including T- and B cells, plasma cells, and NK-cells (Figure 1B-D). The cluster of cycling cells could be segregated into myeloid cell and T cells (Figure 1E). Overall, we could identify a wide range of synovial cell types and their subpopulations, confirming the heterogeneity of ST.

**Figure 1.**
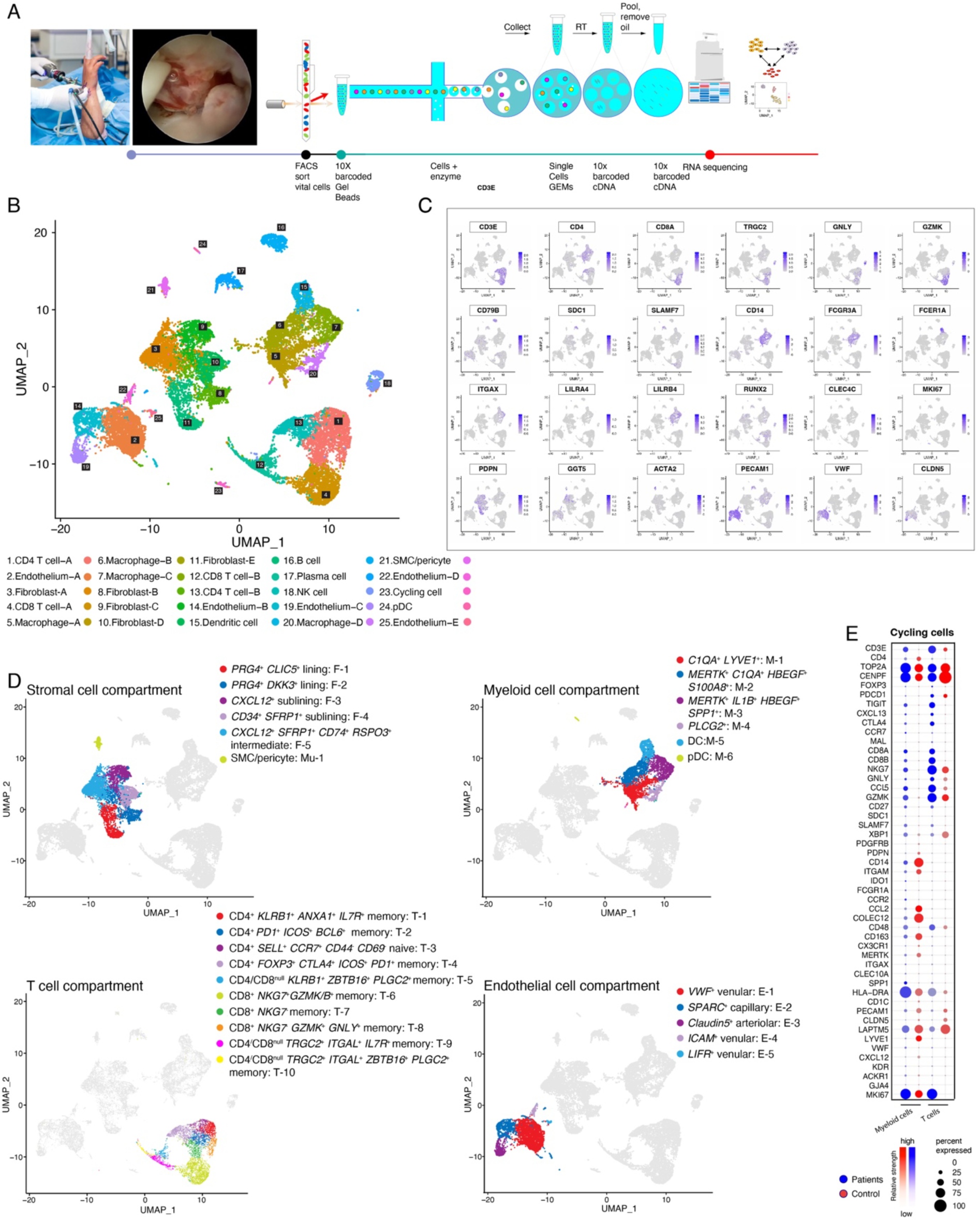
Unsupervised clustering of scRNA-seq data. (A) Schematic overview illustrating the synovial tissue sampling and processing to obtain high quality single-cell RNA seq data. (B) UMAP plot showing 25 distinct cell clusters. (C) UMAP plots showing normalized expression of canonical marker genes. (D) UMAP plots showing cell subtypes within main lineages. (E) Dotplots of cycling cells.

### Alterations in cellular composition of ST in RA versus control

Next, we set out to compare the cellular composition of the ST of RA patients with the control sample. For the patients and control, we analyzed a total of 17,176 and 7,884 high-quality cells, respectively (Figure 2A). In the two RA samples combined, we identified cluster abundances within the major cell lineages, including T cells (n=6,229 cells), myeloid cells (n=4,351 cells), endothelial cells (n=2,256 cells), fibroblasts (n=2,156 cells), B- and plasma cells (n=1,374 cells), and NK cells (n=522 cells). Despite overlapping profiles, both RA patients displayed distinctive characteristics in cellular composition. In the control subject, we observed clusters of T cells (n=324 cells), fibroblasts (n=4,490 cells), endothelial cells (n=1,806 cells), and myeloid subsets (n=1,077 cells), but only few cells that made up the compartments of B cell/plasma cell (n=4 cells) and NK cells (n=26 cells). These findings were confirmed by histology(Figure 2B).

**Figure 2.**
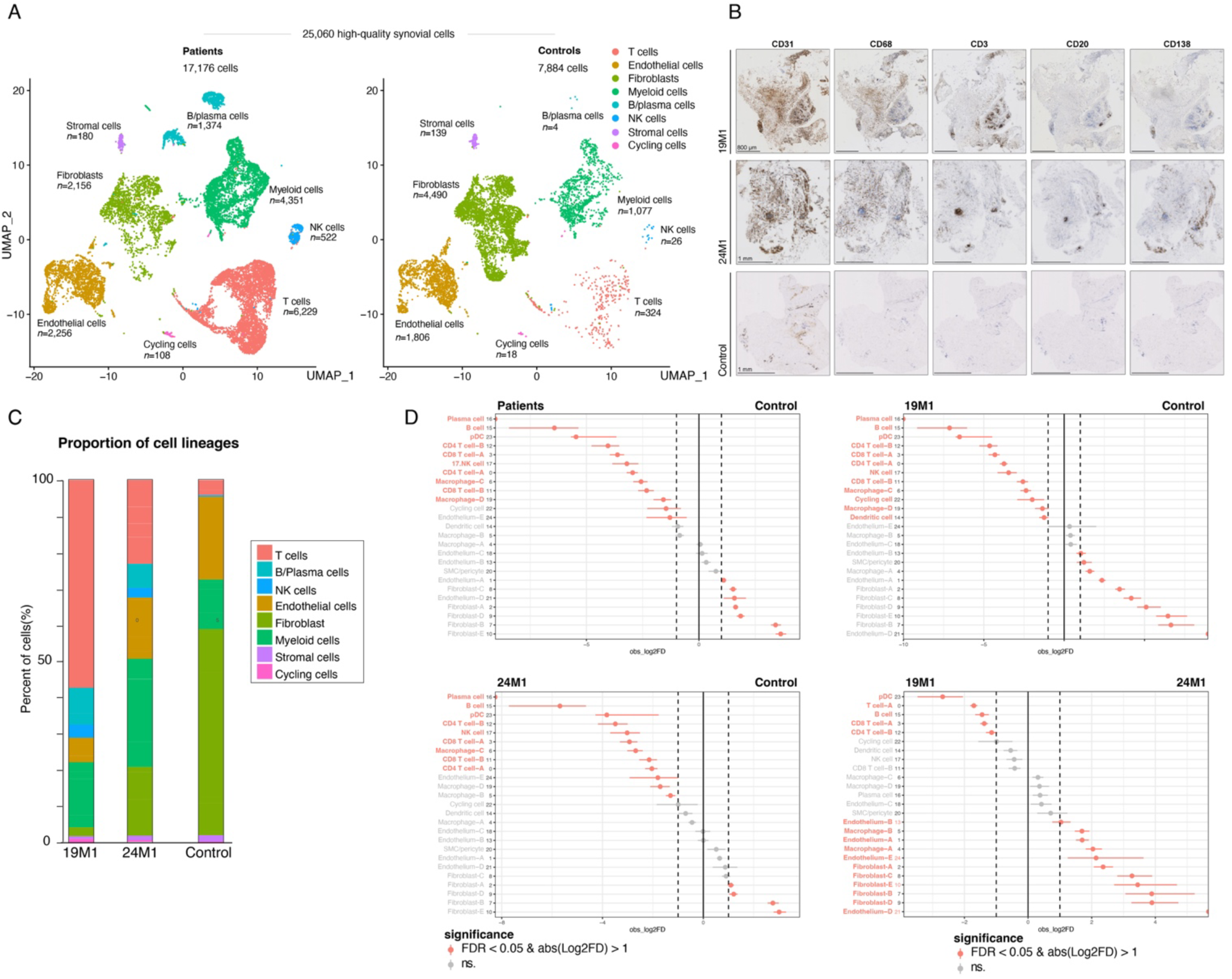
Alterations in cellular composition of ST in RA versus control (A) UMAP plots showing cell clusters for RA patients or control subject. (B) Representative histology images of the ST fragments from 19M1 (top row), 24M1 (middle row), and control (bottom row); immunostaining for CD31, CD68, CD3, CD20, and CD138. (C) Proportion of cell lineages for each sample. (D) Relative differences in cell proportions for each cluster between RA patients versus control. Red clusters have an FDR < 0.05 and mean log2 fold enrichment >1 (permutation test; n=10,000).

In line with previous reports, ST derived from the RA patients showed proportional increases of the compartments of T cell subsets, B- and plasma cells, myeloid populations, and NK cells, as compared to that of the control (Figure 2A,B). This was confirmed by a permutation test that calculates the relative differences in cluster proportions between RA patients and control (Figure 2C). Remarkedly, when we compared patient 19M1 to 24M1, we found that the cell clusters CD4 T cell-A and pDC were enriched in patient 19M1, and in contrast, proportions of the clusters of fibroblasts and endothelium cells were proportionally increased in patient 24M1 (Figure 2D). Although, in general, we could detect relevant differences in cellular composition between RA and control, the more compelling feature is the presence of interpatient variability. This variability may determine the relationship between RA synovial signature and the clinical phenotype, potentially influencing treatment response and outcome.

### Cellular and molecular pathways in RA ST identified by scRNA-seq analysis

To further characterize the ST transcriptome in RA, we performed functional enrichment analysis on cell type-independent transcripts that were differentially expressed in patients, as compared to control. In line with our cellular composition data, gene ontology (GO) enrichment analysis to the domains of biological process and molecular function revealed pathways of adaptive immunity involving T- and B cell receptor (TCR; BCR) signaling pathways, leukocyte cell-cell adhesion, and major histocompatability (MHC) class II signaling (Figure 3A), referring to ectopic germinal center formation, known to RA ST[19]. Coincidentally, we noticed enrichment of pathways involving protein synthesis, immune cell energy metabolism, and resource allocation [20]. Pathways that were downregulated involved, amongst others, cell migration, cell-matrix adhesion, and integrin pathways, possibly hinting at interference with immune cell trafficking from the inflammatory site (Figure 3A).

**Figure 3.**
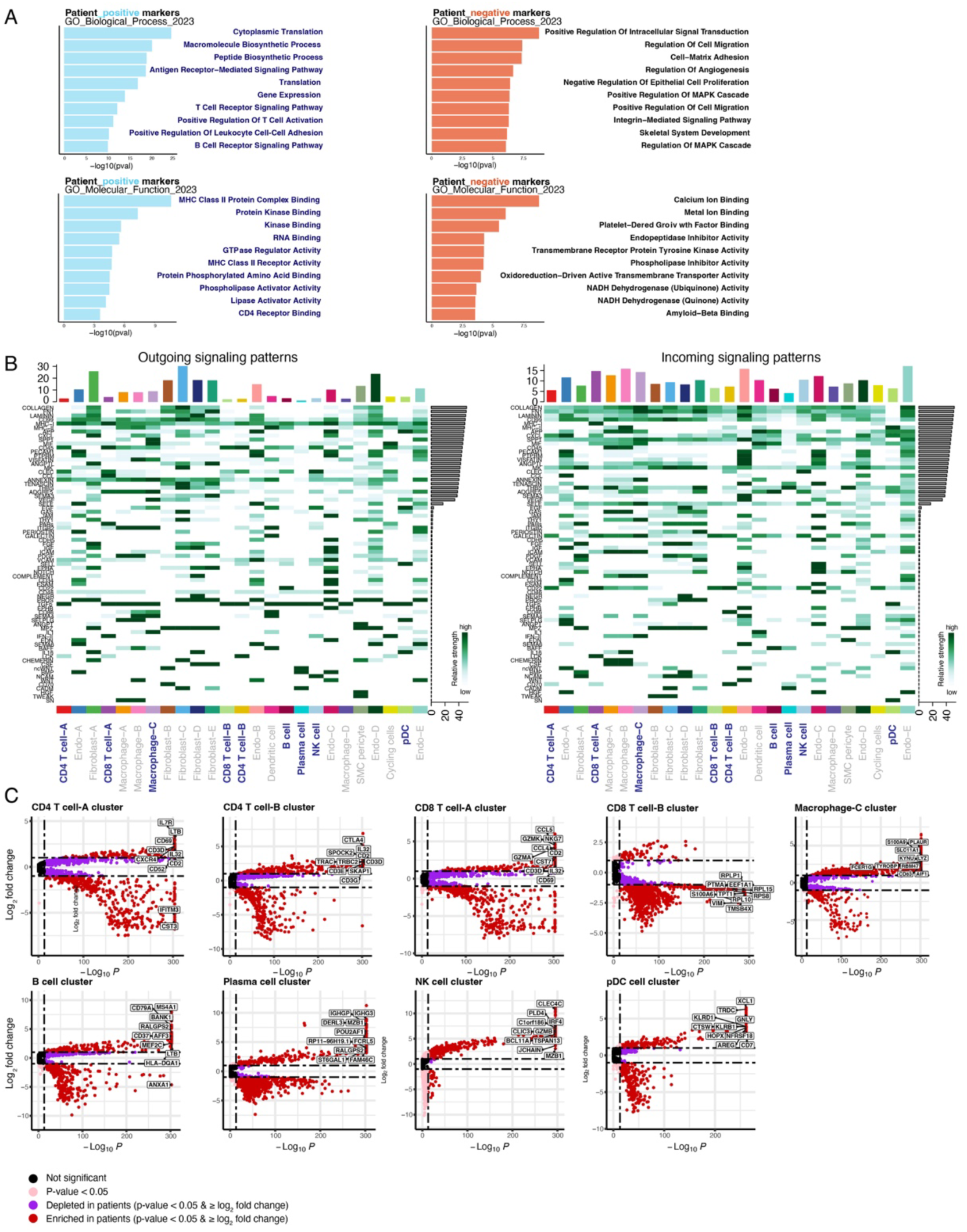
Cellular and molecular pathways in RA ST identified by scRNA-seq analysis (A) Functional enrichment analysis of DEGs of RA ST compared to control. (B) CellChat heatmap showing the relative strength of outgoing and incoming signaling pathways for each cluster in RA compared to control. Vertical bars on top of the heatmap indicate overall importance of the cluster in the respective signaling pattern (i.e. outgoing or incoming). Horizonal bars indicate overall importance of the signaling pathway in the respective signaling pattern. (C) Volcano plots showing distinct DEGs in RA vs control between indicated cluster and all other clusters.

Cellchat[17] allowed us to analyze the intercellular crosstalk within RA ST (Figure 3B). Amongst the top 25 of enriched output patterns, pathways that stood out in CD4 and CD8 T cell subsets and NK cells related to polarization/activation, chemotaxis, cytokine production, and response to corticosteroids, whereas B cells showed enrichment of pathways that related to cytokine production, BCR signaling and autoimmunity. The outgoing communication of myeloid subsets had a broad impact on many pathogenic mechanisms, including chemotaxis, vascular homeostasis/remodeling, ECM interactions, and cytokine-mediated signaling pathways. Further, pDCs were focussed on transendothelial migration, T cell activation, vascular homeostasis/remodeling, and joint surface defect repair/homeostasis. Interestingly, the CD45- and TGF-ß-mediated pathways seemed to contribute significantly to outgoing communication, given their critical effect on T cell function/MHC signaling, and cell cycling, respectively. For some synovial populations, we noticed overlap between incoming and outgoing signaling patterns.

Next, we further zoomed in on the top 10 of DEGs that discriminated individual aberrant clusters from other clusters across both conditions (i.e. “patient” and “control”) (Figure 3C). In RA, the B cell cluster displayed upregulation of genes associated with BCR signaling, activation and proliferation. In plasma cells, we noticed enrichment for several genes that related to co- and post-translational modifications of immunoglobulins, retrograde protein transport and ER-To-Cytosol transport. As for pDCs, the expression of T- and NK cell regulating cytotoxicity genes stood out, in addition to genes, that related to antimicrobial activity. The NK cluster engaged in enhanced ion channel and (membrane) transport functions, chromatin remodeling and cell migration and adhesion. Aside from canonical activation markers, the CD4 T cell type-A cluster showed enrichment of genes related to secretion of cytokine-mediated responses and cell migration/adhesion. In the CD8 T cell cluster, we noticed increased expression of genes involved in processing of pro-granzymes into proteolytically active forms. In contrast, the CD4/CD8^null^ T cell cluster, most likely comprising TRGC2^pos^ gammadelta T cells, appeared less active, particularly in terms of differentiation, and migration. Finally, a large set of effector genes in the Macrophage-C subset, related to secretory granules/pathways, defense response to bacteria, and myeloid cell activation [21, 22].

In summary, these analyses revealed enrichment of cell type-independent pathways that are consistent with previous ST studies[10], and at a celltype-specific level may provide criticial and novel information on cellular and molecular mechanisms that may add to the existing pool of knowledge of the pathogenesis of RA.

### Generic communication patterns of the major cell lineages within RA synovial tissue

Understanding the cellular interactions within ST is crucial for identifying appropriate therapeutic targets. Hence, we searched for generic communication patterns, divided into outgoing and incoming signals. Using Cellchat, we could identify 4 generic communication patterns, that related to the (1) immune cell compartment, (2) myeloid cell compartment, (3) fibroblast populations, and (4) endothelial cell clusters. Outgoing pathways that stood out in the immune cell compartment were associated with interferon-gamma mediated pathways, cell adhesion, T cell polarization/stimulation, MHC signaling, BCR signaling, autoimmunity, and cytokine production (Figure 4A). Knowing their capacity to evolve into a highly inflammatory state[15], the fibroblast subpopulations altogether displayed a phenotype that may have impact on cell migration and recruitment and adhesion, vascular homeostasis/remodeling, cell apoptosis, joint surface defect repair or homeostasis, complement cascade, apoptotic process, cell-cell interactions, inflammatory response, ECM-interaction/binding, and cytokine-mediated signaling pathways (Figure 4A). Myeloid cell clusters showed particular enrichment in output pathways related to metabolism, stress response, T cell polarization/stimulation, B cell survival, and cytokine-mediated signaling (Figure 4A). The endothelial cell compartment coordinated outgoing signals that had impact on cell-cell adhesion, leukocyte transendothelial migration, and maintenance of vascular permeability/cell-cell junction. A noticeable overlap between incoming and outgoing communication patterns was observed in the immune cell clusters, where we noted enrichment of several effector/activation markes (Figure 4B).

**Figure 4.**
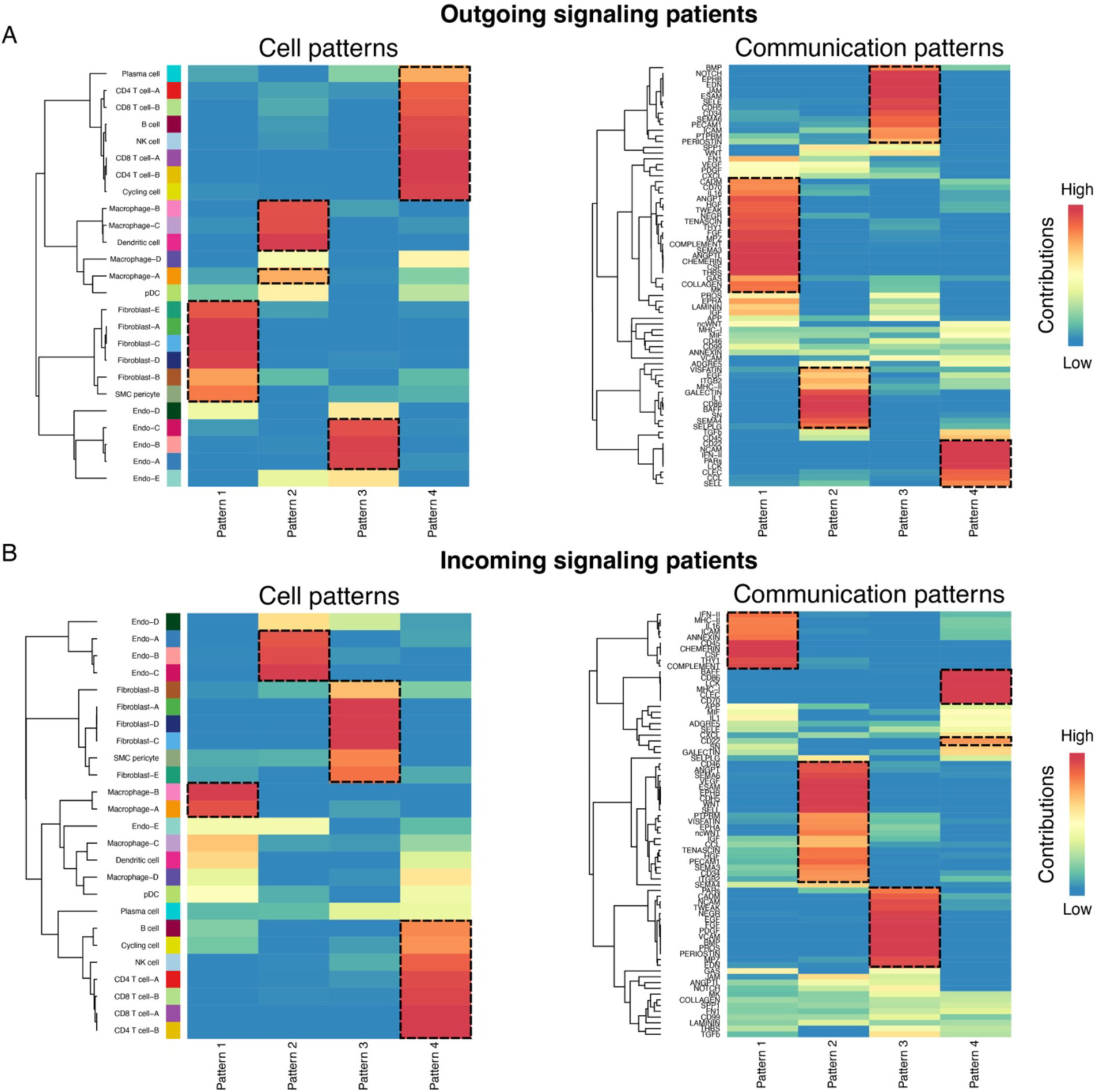
Generic communication patterns of the major cell lineages within RA synovial tissue (A-B) Heatmaps showing cell and communication patterns for (A) outgoing) and (B) incoming signaling in synovial transcriptome of RA patients compared to control. Cell clusters and pathways are hierarchically clustered based on similar contribution. Colors scale shows the probability of contribution.

### Contribution of CTAPs to outgoing and incoming signaling of communication networks

A recent paper, addressing the ST heterogeneity in RA pathogenesis, defined six different cell-type abundance phenotypes (CTAP) based on the relative enrichment of the six major cell lineages, involving leucocyte-poor and pauci- and pleural immune cell phenotypes[10]. These CTAPs proved to be versatile and subjected to disease state and treatment, and, more relevant, had clinical utility to potentially predict treatment response[10]. Using this methodology, we tested the frequency of the different CTAPs in our dataset, and found that the T cell/myeloid cell (TM) CTAP predominated in patient 19M1 (75%), whereas the ST of patient 24M1 was enriched for the endothelial cell/fibroblast/myeloid cell (EFM) CTAP (67%) (Figure 5A).

**Figure 5.**
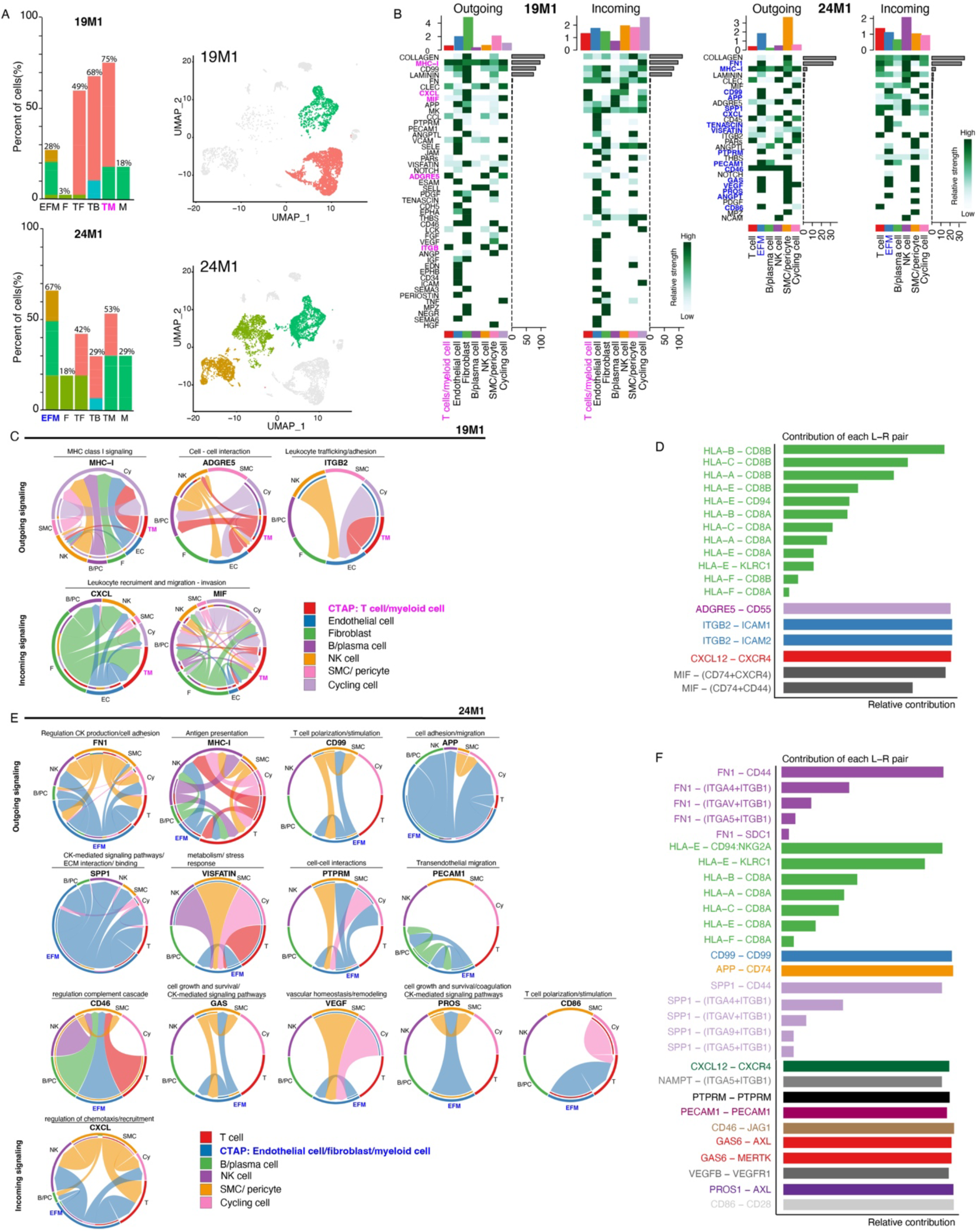
Contribution of CTAPs to outgoing and incoming signaling of communication networks (A) The percentage of cells that could be ascribed to one of the six CTAPs in each RA patient. EFM, endothelial fibroblast and myeloid cells; F, fibroblasts and the pauci- and pleural immune phenotypes; TF, T cells and fibroblasts; TB, T and B cells; TM, T cells and myeloid cells; M, myeloid cells. The UMAP plots highlight the cell clusters that are part of the most prevalent CTAPs. (B) Heatmaps showing outgoing and incoming signalling for every CTAP in each patient. Vertical bars on top of the heatmap indicate overall importance of the cluster in the respective signaling pattern (i.e. outgoing or incoming). Horizonal bars indicate overall importance of the signaling pathway in the respective signaling pattern. (C and E) Chord diagrams showing the communication score between interacting cell clusters for distinct signaling pathways. Width of edge represent the interaction strength (D and F) Barplots showing the relative contribution of ligand-receptor pairs within the indicated pathway interactions in C and E.

Next, we investigated the relative contribution of the CTAPs to the outgoing and incoming signaling of communication networks. Cellchat analysis in patient 19M1 (Figure 5B), inferred that the TM CTAP was enriched in pathways related to chemotaxis/recruitment, antigen presentation, and inflammatory response. In this regard, the TM CTAP seemed to interact with the cell compartments of endothelial cells, B- and plasma cells, and cycling cells (Figure 5B). When we looked at CTAP TM as receiver population, Cellchat predicted interactions, mostly with the compartments of fibroblasts, about regulation of chemotaxis/recruitment and macrophage function (Figure 5C,D). In this respect, several important ligand-receptor pairs were predicted to be relevant, as shown in (Figure 5C,D).

In patient 24M1, Cellchat analysis inferred that the sender and receiver signaling patterns of the CTAP EFM were distributed across 16 different signaling pathways (Figure 5E). Amongst others, these predicted interactions related to pathways involved in cytokine-mediated signaling and production, T cell polarization and stimulation, cell recruitment, ECM-interaction and binding, complement cascade, and vascular homeostasis/remodeling. We classified these groups of signaling pathways based on structural and functional similarity (Figure 5E). In this respect, several significant signaling ligand-receptor pairs that could be relevant were identified (Figure 5F). In summary, these data provide insights into what extent these CTAPs may be utilized to determine disease state and enable clinical utility prediction of treatment response.

### Module scores related to treatment responses in CTAP and other constituents of ST

Given that CTAPs may be used to predict the effect of therapy [10], we looked per individual patient at the composite expression of druggable gene targets, obtained from previously reported canonical markers, that jointly (i.e. as a module) may predict treatment responses to currently available and future classes of DMARDs for RA (Figure 6A). For this, we aimed to identify signature gene sets or module scores that are either expressed in the ST as a whole or more specific to the prevailing CTAP, relative to other cell lineages. For the RB19 and RB24 datasets, we extracted 20 modules scores, and these turned out not to be mutually exclusive (Figure 6A). In the ST of patient 19M1, we found enrichment for the module scores related to the JAK and CXCR4 pathways, and to lesser extent the glucocorticosteroid response (NR3C1). It was noticed that the prevailing CTAP TM appeared to contribute substantially to these enriched module scores (Figure 6B). Notably, we found enrichment of a wider spectrum of druggable gene targets in patient 24M1, as compared to 19M1, including module scores related to JAK, CXCR4, ITGA4, and NR3C1 pathways and to lesser extent TNF (Figure 6A). It appeared that the prevailing CTAP EFM contributed in a substantial manner to the enrichment of druggable gene targets related to the JAK pathway, glucocorticosteroid response, and CXCR4, however also the T cell subsets, and cycling cells had a contributive role in regard to CXCR4, ITGA4, and glucocorticosteroid response (Figure 6B).

**Figure 6.**
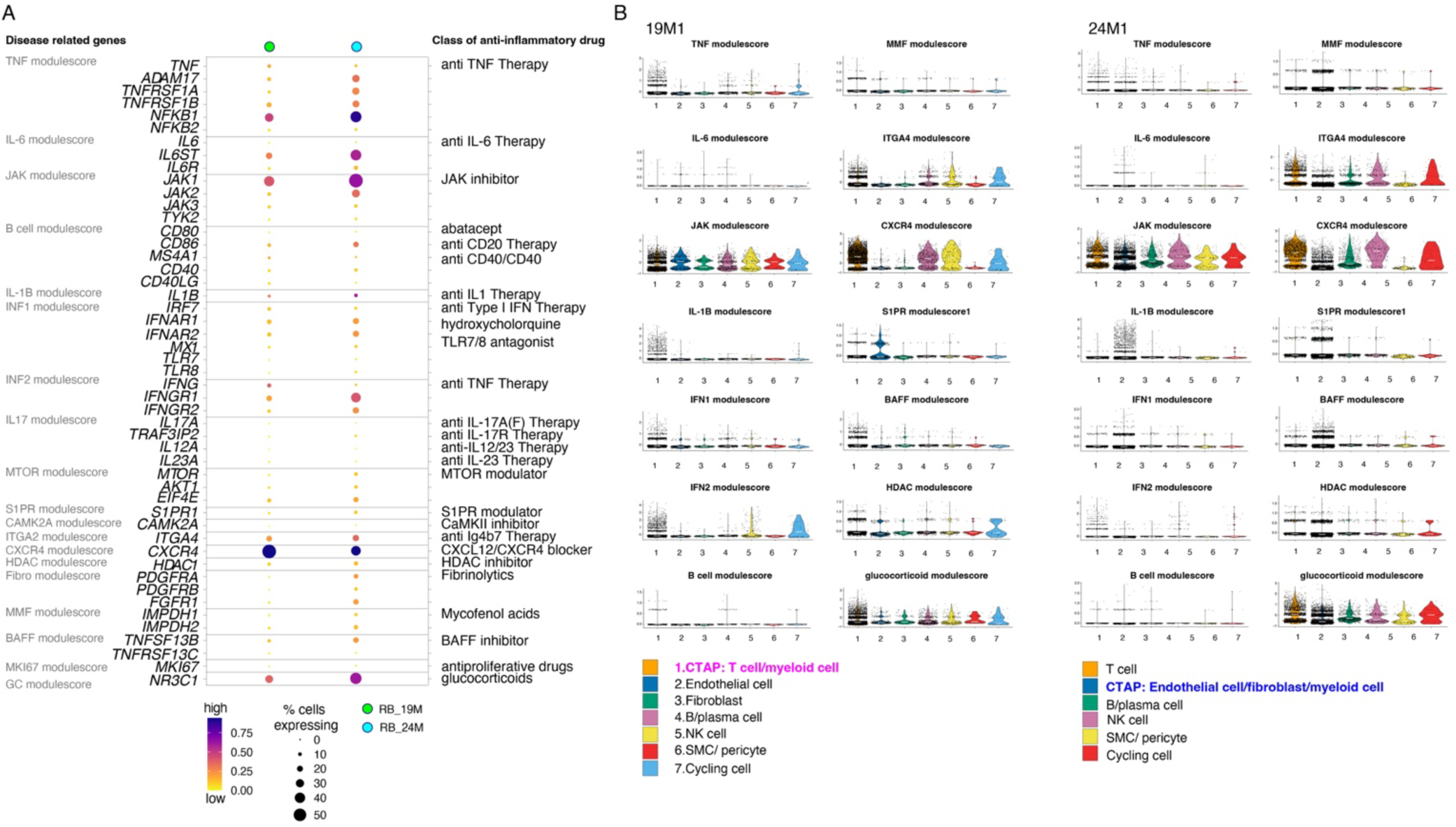
Module scores related to treatment responses in CTAP and other constituents of ST (A) Dotplot showing the normalized expression of the module scores of the druggable gene targets. (B) Violin plots showing the contribution of the CTAP to the enrichment of druggable gene targets.

## DISCUSSION

In this exciting era, omics technologies like scRNA-seq enable integrative and systematic profiling of cellular heterogeneity, molecular functions, and communication networks within target tissues of patients with chronic immune diseases like RA. By using this method, our intentions were to make a reference map of human ST for a better grasp on cell-type specific mechanisms that underlie disease pathogenicity in a singular patient, and to define cellular and molecular landmarks that may help to build a clinical instrument for guiding rheumatologists and immunologists in the selection of specific b- and tsDMARD therapy at an individual level. In this feasibility study, we demonstrate that our approach on ST derived from minimally invasive arthroscopic surgery is successful in retrieving informative and high-quality, single-cell transcriptome data. In a future clinical setting, patients presenting with wrist arthritis as prominent feature, would undergo therapeutic synovectomy [23], where analysis of their removed ST will provide the rheumatologist with a tailored cellular and molecular roadmap to start systemic treatment.

In line with earlier reports on RA ST [5, 6, 10], our observations, from a macroscopic point of view, show cardinal alterations in the target tissue of RA, as compared to control synovium, involving infiltration of effector immune cells that was comprised of T cell subsets, B- and plasma cells, pDCs, and NK cells, in conjunction with augmented proportion of macrophage subsets. Further, generic enrichment analysis of the RA dataset, as a whole, revealed enrichment of processes such as T- and B cell receptor signaling and activation, leukocyte cell-cell adhesion, and MHC class II signaling, suggesting formerly reported cellular events that transpire in highly specialized structures within synovial joints in RA, coined as germinal center response [19]. Individually, the aberrant clusters that emerged, displayed cell-type specific enrichment of pathways that fit their proclaimed role in the immunopathogenesis of RA, and may drive stromal tissue dysregulation to perpetuate a chronic inflammatory state. Building on recent findings, we were able to allocate one of the six CTAPs, as reported recently, to each of our two RA samples[10]. Deeper analysis of the unique CTAP signature gave insights into the communication pathways and networking with other cell lineages, and provided us with predictive gene modules for therapy response, that were not mutually exclusive, and very usefull for weighing the signal intensity of, and pinpointing the therapeutic response to specific constituent parts of the ST.

Interpretation of our omics data reveals that, in addition to how many of which cell clusters are enriched or depleted in the target ST, the number, type, and intensity of cell-cell interactions and signaling pathways are equally important in determining potential druggable gene targets at an individual level. As it appears, transcriptomic differences are not primarily dictated by compositional changes in immigrant immune cell populations, given that cell lineages other than the annotated CTAP may dominate communications patterns (sender and receiver), and propagate the immunopathogenesis, as observed in patient 19M1. In addition, qualitative changes in the stromal compartments may be key drivers of the immunophenotype (see 24M1), and thus may alter the initial therapy response in due course. Altogether, these pieces of information underscore the concept of a heterogeneous disease that share a common arthritic phenotype upon variable activity of diverse pathways, and may be translated into patient-specific ST signature(s) that may serve as stratifier for long-term treatment with b- or tsDMARDs[1].

The concept of selecting the right drug target at the earliest convenience in the individual patient (*’the-earlier-the-better principle’)* has been raised to increase the opportunity of sustained clinical remission, and is underlined by several studies [24]. Others have coined this phenomenon as ‘window of opportunity’, a transient time frame in which the disease is more susceptible to treatment [25]. In this line of thinking, multiple cycles of ineffective DMARDs might select a clinical subtype that is resistant to new treatments. In this regard, a recent study showed that first-line combination of a TNF-inhibitor and MTX yielded better clinical response than step-up treatment with anti-TNF therapy in RA patients that fail to achieve clinical remission with MTX [26]. In the BeSt study, where 4 treatment strategies were compared, MTX step-up therapy with a TNF-inhibitor or a step-up schedule with additional csDMARDs resulted in slower restoration of functional impairment and increased radiologic damage in the first year, as compared to “aggressive” first-line strategies combining a TNF-inhitor or high prednisolone with MTX [27]. These data suggest a change in biology, where a refractory state results from a reset of the balance between pro- and anti-inflammatory cytokine networks in the synovium upon (delayed) iatrogenic depletion of predominant cytokines, or changes in the epigenetic landscape or ‘tissue programming’ (chromatic accessibility at specific genomic loci)[2]. In other words, early effective treatment may prevent progression to therapy refractory disease[2].

Amongst the limitations of our study are high costs and complex logistics related to the scRNA-seq method, which culminated in a small sample size. Also, our interpretation of the intercellular crosstalk and communication patterns are largely based on in-silico based assumptions. Notwithstanding, we were able to delineate a high degree of synovial transcriptomic heterogeneity, which paves the way for a follow-up study with a large set of participants where we can validate these data and determine its relationship with the clinical phenotype, therapy response and evolution of RA over the individual’s lifetime. It is anticipated that, over time, not only the sequencing costs will be reduced, but also computational frameworks to handle large dataset will become more accessible. Ultimately, this will benefit the scalability of scRNA-seq and its wide implementation. In our perception, there are several requirements before implementation in daily practice can be realized: (1) procedures for obtaining ST need to be low-risk for the patient and joint arthroscopy needs to be accessible for the rheumatologist, (2) biopsy and tissue analysis needs to be performed timely to prevent long periods of undertreatment, and (3) test accuracy needs to be high to ensure reliable predictions [2, 28, 29]. Except for a timely analysis, all of these requirements were met in this feasibility study, and thus merits further testing in a larger set of patients with RA.

In conclusion, we demonstrated that ST from RA patients can safely be obtained from the wrist by means of arthroscopy and is ample for cellular studies. Secondly, we showed that our scRNA-seq approach successfully reveals the heterogeneity of ST by identifying a wide range of synovial cell types and their subpopulations, as well as relevant enrichment of cellular and molecular signatures that could discriminate between RA and control tissues. Additionally, we observed interpatient variability that may ultimately enable us to determine the relationship between RA synovial signature, clinical phenotype, and treatment response/outcome. Previously reported CTAPs were present and could be related to specific molecular signatures for therapy response, which were not mutual exclusive. Our data pave the way for evaluation of this approach in follow-up studies with a large set of participants, enhancing the robustness and generalizability of the data in order to develop an instrument for driven/stratified approaches to treating patients with RA.

## Data Availability

All data produced in the present study are available upon reasonable request to the authors

Raw sequencing data have been deposited in the NCBI GEO repository (GSE number follows) and will be publicly available as of the date of publication. The code used for analysis is available upon request.

## ACKNOWLEDGEMENT

We thank C. van Helden-Meeuwsen (Immunology department, Erasmus MC, Rotterdam, Netherlands) for technical support.

